# The introduction of standardised packaging and availability of illicit cigarettes: a difference-in-difference analysis of European Union survey data 2015-2018

**DOI:** 10.1101/2020.08.26.20182329

**Authors:** Anthony A Laverty, Christopher Millett, Nicholas S Hopkinson, Filippos T Filippidis

## Abstract

Standardised packaging of tobacco products is intended to reduce the appeal of smoking, but the tobacco industry claims this increases illicit trade. We examined the percentage of people reporting being regularly offered illicit cigarettes before and after implementation of standardised packaging in the United Kingdom, Ireland and France and compared this to other European Union countries. Reported regular illicit tobacco exposure fell from 2.7% to 2.3% between 2015 and 2018 in the three countries implementing the policy, and from 2.3% to 1.7% in control countries (p for difference=0.320). Standardised packaging does not appear to increase the availability of illicit cigarettes.

## INTRODUCTION

In 2012 Australia became the first country to require standardised (or plain) packaging for tobacco products. The tobacco industry argued that this strategy is ineffective (1) and will increase illicit tobacco use. There is limited independent evidence on this issue, although the UK Chantler review concluded that there was no evidence of an increase in illicit trade in Australia after implementation (2). The recent World Trade Organization decision that standardised packaging is consistent with international trade law will encourage other countries to implement this policy, but also increase misinformation and lobbying by the tobacco industry (3). The aim of this study is to assess whether the implementation of plain packaging in the UK, France and Ireland was associated with a change in the likelihood of being offered illicit cigarettes compared with other European countries.

## METHODS

We analysed individual-level data from waves 84.4 (collected November/December 2015; n=27,672) and 90.4 (December 2018; n=27,636) of the Special Eurobarometer Survey (4)(5). Standardised packaging has been in force for tobacco sold in France from January 2017, in the UK from May 2017, and for tobacco manufactured after 30^th^ September 2017 in the Republic of Ireland.

The outcome was reporting being offered illicit cigarettes, assessed with the question: “Have you ever been offered black market cigarettes to buy or smoke?”. Responses were “No, never”; “Yes, rarely (< once a month)”; “Yes, occasionally (1-3 times per month)”; and “Yes, frequently (once per week or more)”.

Socio-demographic data included age (15-24; 25-39; 40-54; 55+ years); sex; residence type (rural, town/suburb or city); age at completion of education (0-15 years; 16-19 years; 20+ years; and still studying); occupation (employed; unemployed); smoking status (non-smoker, current smoker and former smoker); and difficulty paying bills during the last year (almost never/rarely; occasionally; or most of the time).

We extracted country-level data on Gross Domestic Product (GDP) per capita from Eurostat and the Corruption Perception Index (CPI) from Transparency International, as per our previous work (6). Tobacco Control Scale scores (https://www.tobaccocontrolscale.org/) were used to capture national tobacco control policies. We excluded the price element of the score and used Weighted Average Price of cigarettes (WAP) provided by the European Commission and adjusted these for inflation using the Harmonised Index of Consumer Prices (HICP) (https://ec.europa.eu/eurostat).

We employed a difference-in-difference approach using a two-level ordered (random intercept) regression model. The two-level model accounts for clustering of individuals’ responses (first level) within countries (second level).

In the difference-in-difference model, we created a binary variable (countries that introduced plain packaging vs. countries that did not), a year variable (2015 vs 2018) and an interaction term between these two (the difference-in-difference estimate). The model included CPI, GDP per capita, and WAP (country level); age, sex, residence type, age when completed education, occupation, smoking status and difficulty paying bills (individual level). The three countries which implemented standardised packaging (intervention) were compared with the other 25 EU countries (control). We also ran ordered regression models in each country adjusting for the same factors.

Results are presented as population weighted mean and adjusted Odds Ratios (aOR) with 95% Confidence Interval [CI] of reporting a higher level of exposure to illicit tobacco. Ordered regression uses all of the responses to the outcome question and gives odds for being offered illicit cigarettes more often. Odds ratios above one represent an increased frequency of being offered, and vice versa. Sensitivity analyses were performed using data from smokers only, and excluding countries with land borders with non-EEA countries, which has previously been linked to greater illicit cigarettes availability (6).

## RESULTS

In the three intervention countries 7.1% (CI 6.0%; 8.3%) were offered illicit cigarettes ‘occasionally or regularly’ and 2.7% (2.0%; 3.5%) ‘regularly’ in 2015. In 2018, 6.5% (5.3% to 7.9%) were offered illicit cigarettes ‘occasionally or regularly’ and 2.3% (1.6%; 3.2%) ‘regularly’ (Figure 1).

**Figure 1.**
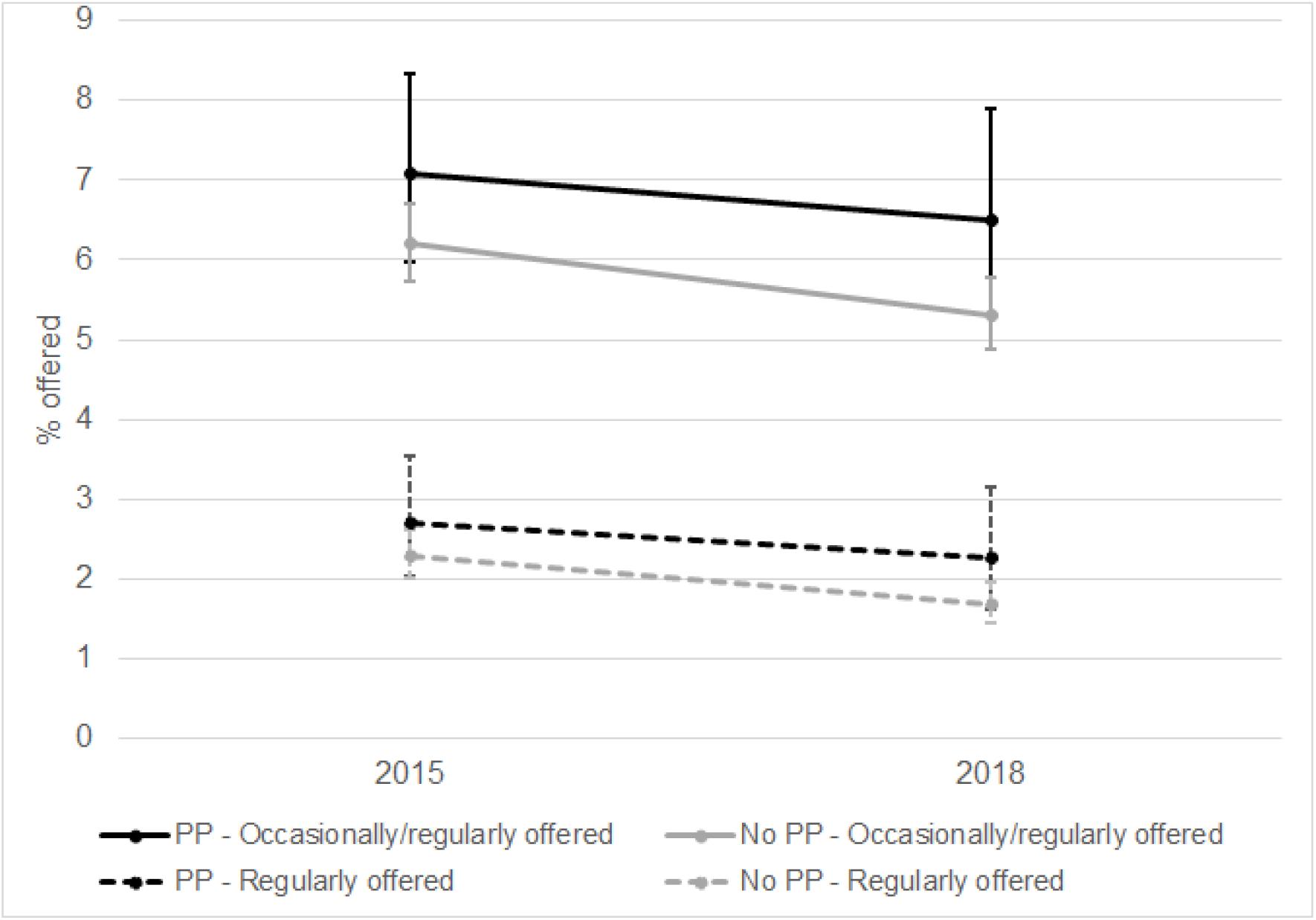
Weighted prevalence of having been offered illicit cigarettes in 2015 and 201S in countries which introduced plain packaging and control countries

In the 25 control countries, 6.2% (5.7%; 6.7%) of respondents were offered illicit cigarettes ‘occasionally or regularly’ and 2.3% ‘regularly’ (2.0%; 2.6%) in 2015. In 2018, 5.3% (4.9%; 5.8%) of respondents were offered illicit cigarettes ‘occasionally or regularly’ and 1.7% (1.4%; 2.0%) ‘regularly’ (Figure 1).

Although changes from 2015 to 2018 varied across the EU (Supplementary Table 1 & Figure 2), the frequency of being offered illicit cigarettes fell between 2015 and 2018 in both control (aOR 0.92 (0.85;0.99)) and intervention countries (aOR 0.85 (0.73;0.99)). These two estimates were not statistically significantly different (aOR for interaction term: 0.93 (0.80; 1.07; p=0.320)) (Figure 3).

**Figure 2.**
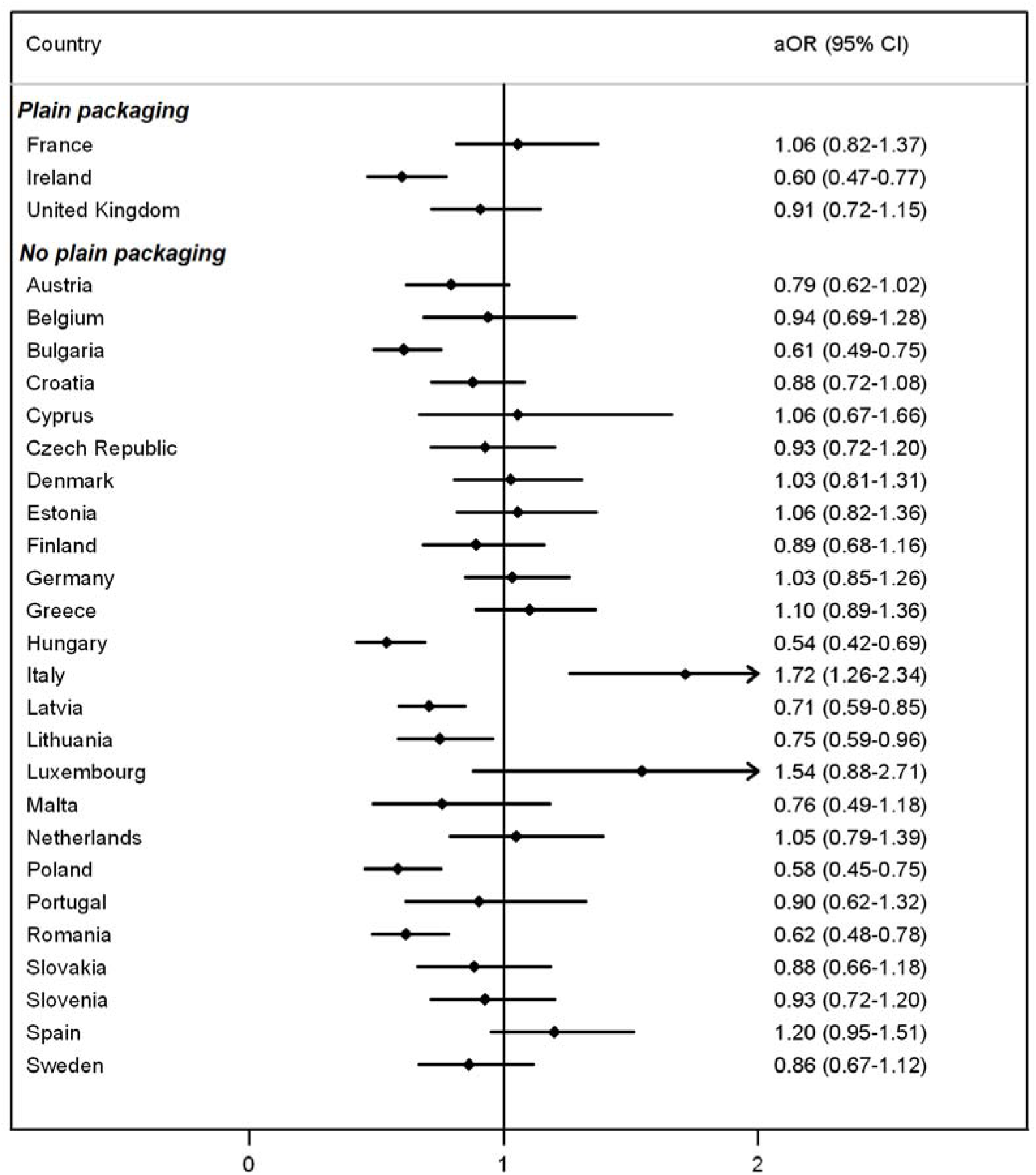
Changes in odds of having been offered illicit cigarettes more frequently between 2015 and 2018, by country.

**Figure 3.**
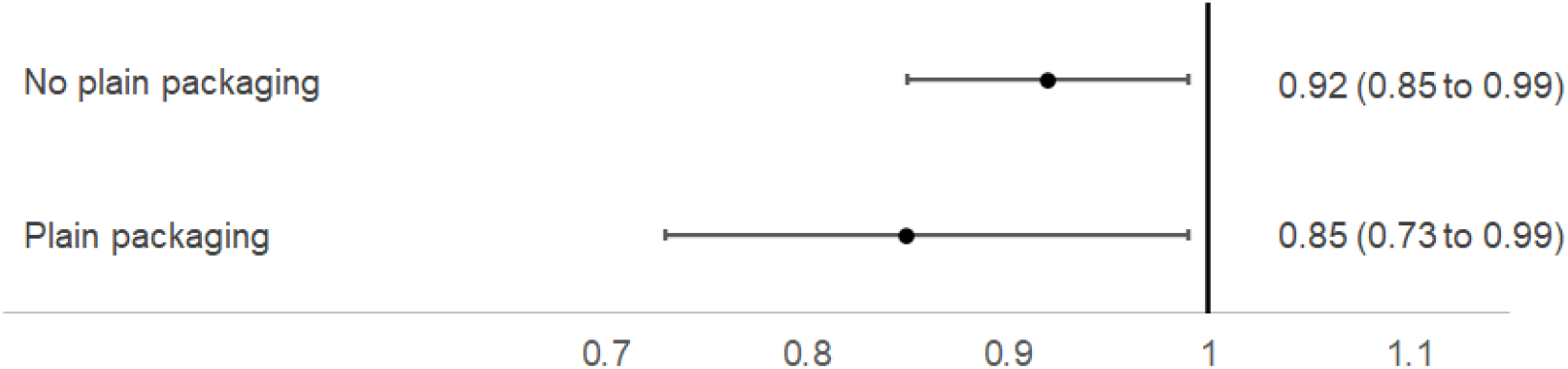
Changes in odds of having been offered illicit cigarettes more frequently between 2015 and 2018 in countries which introduced plain packaging and those which didn’t.

Results for sensitivity analyses among smokers only and among countries with no non-EEA border also found greater increase in the odds of being offered illicit cigarettes more frequently in intervention countries (Supplementary Table 2).

## DISCUSSION

This study is the first to assess whether levels of illicit trade in tobacco have risen after implementation of standardised packaging in Europe. We found no evidence to suggest that this was the case.

This study used nationally representative data from all 28 EU countries and included a range of individual and country level correlates. We used a robust difference-in-difference study design with consistent outcome questions over time and between countries. Nonetheless, rather than the more accepted term “illicit cigarettes” the survey question referred to “black market” cigarettes and this may mean different things in different contexts and locations. The meaning of the term ‘black market cigarettes’ may differ between countries, thus cross-country comparisons of absolute percentages may not fully reflect the availability of illicit cigarettes. However, our analysis compared within country changes in a 3-year period, hence such differences should have minimal effects on our findings. Our analyses are based on self-reported data, and do not provide any insight regarding the actual market share of illicit trade. Nonetheless, Eurobarometer uses a consistent and representative study design and provides unique data on illicit trade, a topic known to be difficult to evaluate (7)(8).

### Conclusions

These results suggest that standardised packaging does not lead to an increase in illicit tobacco. Governments around the world should ignore tobacco industry arguments against standardised packaging and implement this effective policy more widely.

## Data Availability

Eurobarometer data can be downloaded by researchers from https://www.gesis.org/en/eurobarometer-data-service/search-data-access/data-access

---

**Supplementary Table 1.**
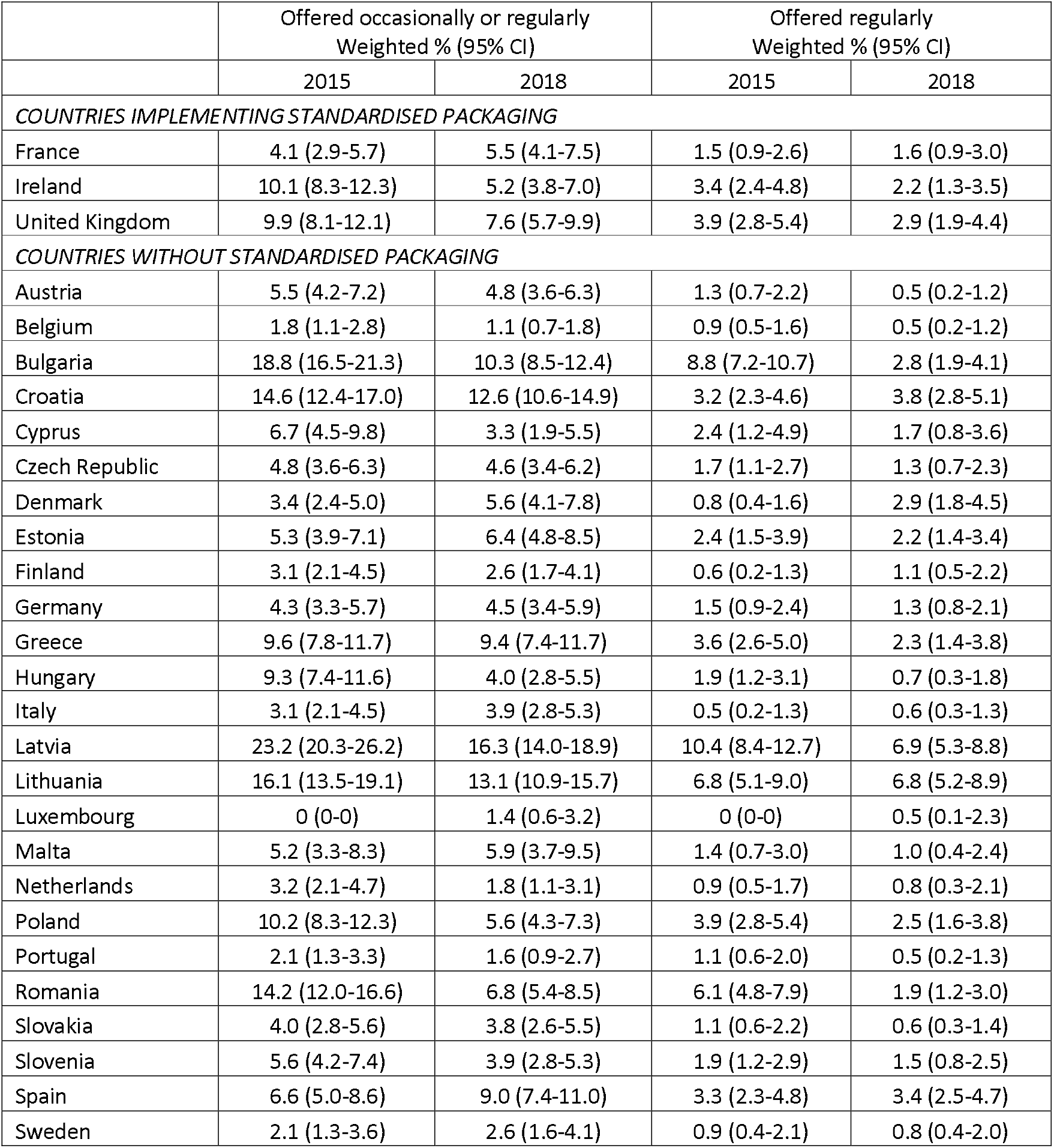
Percentages of reporting being offered illicit cigarettes by individual country and year

|  | Offered occasionally or regularly<br>Weighted % (95% CI) |  | Offered regularly<br>Weighted % (95% CI) |  |
| --- | --- | --- | --- | --- |
|  | 2015 | 2018 | 2015 | 2018 |
| *COUNTRIES IMPLEMENTING STANDARDISED PACKAGING* |  |  |  |  |
| France | 4.1 (2.9-5.7) | 5.5 (4.1-7.5) | 1.5 (0.9-2.6) | 1.6 (0.9-3.0) |
| Ireland | 10.1 (8.3-12.3) | 5.2 (3.8-7.0) | 3.4 (2.4-4.8) | 2.2 (1.3-3.5) |
| United Kingdom | 9.9 (8.1-12.1) | 7.6 (5.7-9.9) | 3.9 (2.8-5.4) | 2.9 (1.9-4.4) |
| *COUNTRIES WITHOUT STANDARDISED PACKAGING* |  |  |  |  |
| Austria | 5.5 (4.2-7.2) | 4.8 (3.6-6.3) | 1.3 (0.7-2.2) | 0.5 (0.2-1.2) |
| Belgium | 1.8 (1.1-2.8) | 1.1 (0.7-1.8) | 0.9 (0.5-1.6) | 0.5 (0.2-1.2) |
| Bulgaria | 18.8 (16.5-21.3) | 10.3 (8.5-12.4) | 8.8 (7.2-10.7) | 2.8 (1.9-4.1) |
| Croatia | 14.6 (12.4-17.0) | 12.6 (10.6-14.9) | 3.2 (2.3-4.6) | 3.8 (2.8-5.1) |
| Cyprus | 6.7 (4.5-9.8) | 3.3 (1.9-5.5) | 2.4 (1.2-4.9) | 1.7 (0.8-3.6) |
| Czech Republic | 4.8 (3.6-6.3) | 4.6 (3.4-6.2) | 1.7 (1.1-2.7) | 1.3 (0.7-2.3) |
| Denmark | 3.4 (2.4-5.0) | 5.6 (4.1-7.8) | 0.8 (0.4-1.6) | 2.9 (1.8-4.5) |
| Estonia | 5.3 (3.9-7.1) | 6.4 (4.8-8.5) | 2.4 (1.5-3.9) | 2.2 (1.4-3.4) |
| Finland | 3.1 (2.1-4.5) | 2.6 (1.7-4.1) | 0.6 (0.2-1.3) | 1.1 (0.5-2.2) |
| Germany | 4.3 (3.3-5.7) | 4.5 (3.4-5.9) | 1.5 (0.9-2.4) | 1.3 (0.8-2.1) |
| Greece | 9.6 (7.8-11.7) | 9.4 (7.4-11.7) | 3.6 (2.6-5.0) | 2.3 (1.4-3.8) |
| Hungary | 9.3 (7.4-11.6) | 4.0 (2.8-5.5) | 1.9 (1.2-3.1) | 0.7 (0.3-1.8) |
| Italy | 3.1 (2.1-4.5) | 3.9 (2.8-5.3) | 0.5 (0.2-1.3) | 0.6 (0.3-1.3) |
| Latvia | 23.2 (20.3-26.2) | 16.3 (14.0-18.9) | 10.4 (8.4-12.7) | 6.9 (5.3-8.8) |
| Lithuania | 16.1 (13.5-19.1) | 13.1 (10.9-15.7) | 6.8 (5.1-9.0) | 6.8 (5.2-8.9) |
| Luxembourg | 0 (0-0) | 1.4 (0.6-3.2) | 0 (0-0) | 0.5 (0.1-2.3) |
| Malta | 5.2 (3.3-8.3) | 5.9 (3.7-9.5) | 1.4 (0.7-3.0) | 1.0 (0.4-2.4) |
| Netherlands | 3.2 (2.1-4.7) | 1.8 (1.1-3.1) | 0.9 (0.5-1.7) | 0.8 (0.3-2.1) |
| Poland | 10.2 (8.3-12.3) | 5.6 (4.3-7.3) | 3.9 (2.8-5.4) | 2.5 (1.6-3.8) |
| Portugal | 2.1 (1.3-3.3) | 1.6 (0.9-2.7) | 1.1 (0.6-2.0) | 0.5 (0.2-1.3) |
| Romania | 14.2 (12.0-16.6) | 6.8 (5.4-8.5) | 6.1 (4.8-7.9) | 1.9 (1.2-3.0) |
| Slovakia | 4.0 (2.8-5.6) | 3.8 (2.6-5.5) | 1.1 (0.6-2.2) | 0.6 (0.3-1.4) |
| Slovenia | 5.6 (4.2-7.4) | 3.9 (2.8-5.3) | 1.9 (1.2-2.9) | 1.5 (0.8-2.5) |
| Spain | 6.6 (5.0-8.6) | 9.0 (7.4-11.0) | 3.3 (2.3-4.8) | 3.4 (2.5-4.7) |
| Sweden | 2.1 (1.3-3.6) | 2.6 (1.6-4.1) | 0.9 (0.4-2.1) | 0.8 (0.4-2.0) |

**Supplementary Table 2.**
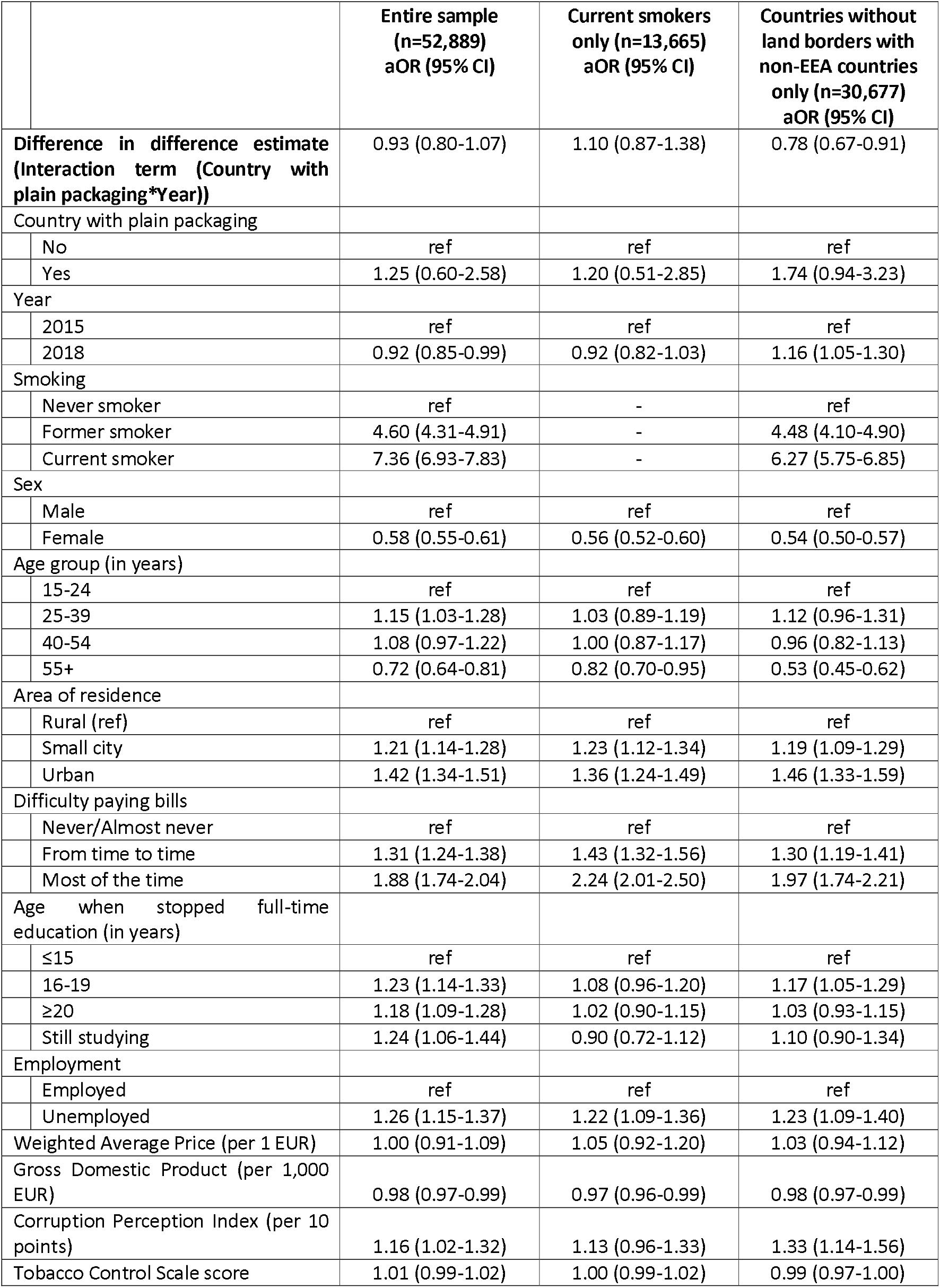
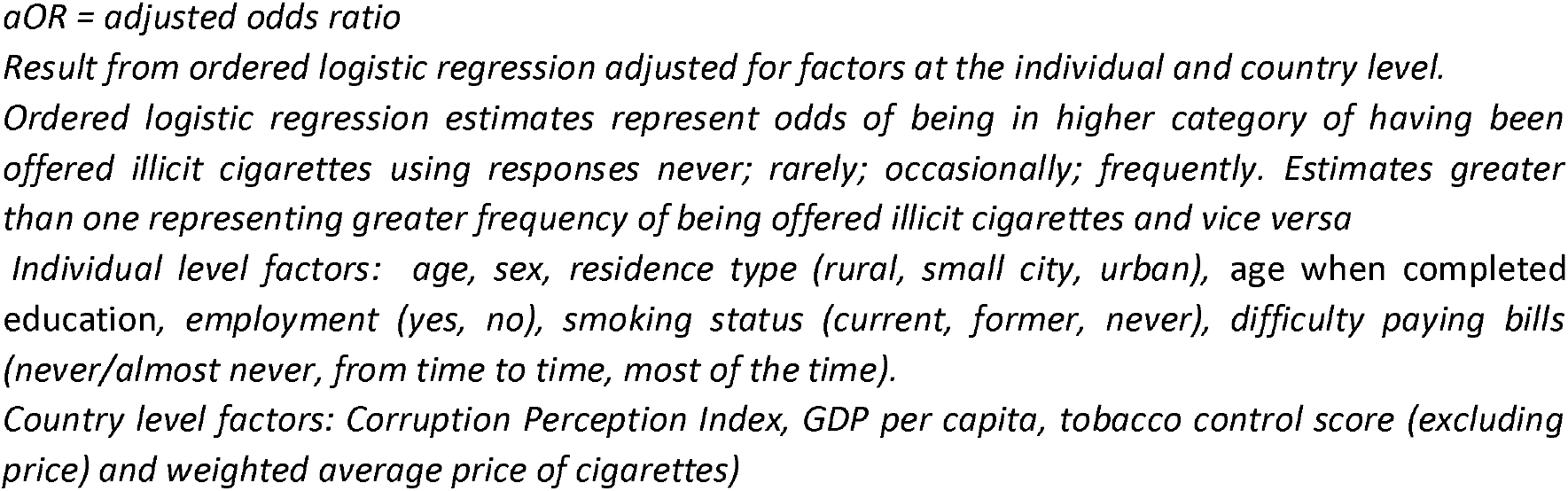
Full regression results and sensitivity analyses from ordered logistic regression of changes in odds of frequency of having been offered illicit cigarettes between 2015 and 2018 in countries which introduced plain packaging and those which didn’t.

|  |  | Entire sample<br>(n=52,889)<br>aOR (95% CI) | Current smokers<br>only (n=13,665)<br>aOR (95% CI) | Countries without<br>land borders with<br>non-EEA countries<br>only (n=30,677)<br>aOR (95% CI) |
| --- | --- | --- | --- | --- |
| **Difference in difference estimate**<br>**(Interaction term (Country with**<br>**plain packaging\*Year))** |  | 0.93 (0.80-1.07) | 1.10 (0.87-1.38) | 0.78 (0.67-0.91) |
| Country with plain packaging |  |  |  |  |
|  | No | ref | ref | ref |
|  | Yes | 1.25 (0.60-2.58) | 1.20 (0.51-2.85) | 1.74 (0.94-3.23) |
| Year |  |  |  |  |
|  | 2015 | ref | ref | ref |
|  | 2018 | 0.92 (0.85-0.99) | 0.92 (0.82-1.03) | 1.16 (1.05-1.30) |
| Smoking |  |  |  |  |
|  | Never smoker | ref | - | ref |
|  | Former smoker | 4.60 (4.31-4.91) | - | 4.48 (4.10-4.90) |
|  | Current smoker | 7.36 (6.93-7.83) | - | 6.27 (5.75-6.85) |
| Sex |  |  |  |  |
|  | Male | ref | ref | ref |
|  | Female | 0.58 (0.55-0.61) | 0.56 (0.52-0.60) | 0.54 (0.50-0.57) |
| Age group (in years) |  |  |  |  |
|  | 15-24 | ref | ref | ref |
|  | 25-39 | 1.15 (1.03-1.28) | 1.03 (0.89-1.19) | 1.12 (0.96-1.31) |
|  | 40-54 | 1.08 (0.97-1.22) | 1.00 (0.87-1.17) | 0.96 (0.82-1.13) |
|  | 55+ | 0.72 (0.64-0.81) | 0.82 (0.70-0.95) | 0.53 (0.45-0.62) |
| Area of residence |  |  |  |  |
|  | Rural (ref) | ref | ref | ref |
|  | Small city | 1.21 (1.14-1.28) | 1.23 (1.12-1.34) | 1.19 (1.09-1.29) |
|  | Urban | 1.42 (1.34-1.51) | 1.36 (1.24-1.49) | 1.46 (1.33-1.59) |
| Difficulty paying bills |  |  |  |  |
|  | Never/Almost never | ref | ref | ref |
|  | From time to time | 1.31 (1.24-1.38) | 1.43 (1.32-1.56) | 1.30 (1.19-1.41) |
|  | Most of the time | 1.88 (1.74-2.04) | 2.24 (2.01-2.50) | 1.97 (1.74-2.21) |
| Age when stopped full-time<br>education (in years) |  |  |  |  |
|  | ≤15 | ref | ref | ref |
|  | 16-19 | 1.23 (1.14-1.33) | 1.08 (0.96-1.20) | 1.17 (1.05-1.29) |
|  | ≥20 | 1.18 (1.09-1.28) | 1.02 (0.90-1.15) | 1.03 (0.93-1.15) |
|  | Still studying | 1.24 (1.06-1.44) | 0.90 (0.72-1.12) | 1.10 (0.90-1.34) |
| Employment |  |  |  |  |
|  | Employed | ref | ref | ref |
|  | Unemployed | 1.26 (1.15-1.37) | 1.22 (1.09-1.36) | 1.23 (1.09-1.40) |
| Weighted Average Price (per 1 EUR) |  | 1.00 (0.91-1.09) | 1.05 (0.92-1.20) | 1.03 (0.94-1.12) |
| Gross Domestic Product (per 1,000<br>EUR) |  | 0.98 (0.97-0.99) | 0.97 (0.96-0.99) | 0.98 (0.97-0.99) |
| Corruption Perception Index (per 10<br>points) |  | 1.16 (1.02-1.32) | 1.13 (0.96-1.33) | 1.33 (1.14-1.56) |
| Tobacco Control Scale score |  | 1.01 (0.99-1.02) | 1.00 (0.99-1.02) | 0.99 (0.97-1.00) |

*aOR = adjusted odds ratio*
*Result from ordered logistic regression adjusted for factors at the individual and country level.*
*Ordered logistic regression estimates represent odds of being in higher category of having been offered illicit cigarettes using responses never; rarely; occasionally; frequently. Estimates greater than one representing greater frequency of being offered illicit cigarettes and vice versa*
*Individual level factors: age, sex, residence type (rural, small city, urban), age when completed education, employment (yes, no), smoking status (current, former, never), difficulty paying bills (never/almost never, from time to time, most of the time).*
*Country level factors: Corruption Perception Index, GDP per capita, tobacco control score (excluding price) and weighted average price of cigarettes)*

## Notes

**Funding** This study is funded by the National Institute for Health Research (NIHR) School for Public Health Research (Grant Reference Number PD-SPH-2015). The views expressed are those of the author(s) and not necessarily those of the NIHR or the Department of Health and Social Care. The funder had no input in the writing of the manuscript or decision to submit for publication.

### Competing Interest Statement

The authors have no conflicts of interest relevant to the content of this article. NSH is the Chair of Action on Smoking and Health (ASH) UK.

### Funding Statement

This study is funded by the National Institute for Health Research (NIHR) School for Public Health Research (Grant Reference Number PD-SPH-2015). The views expressed are those of the author(s) and not necessarily those of the NIHR or the Department of Health and Social Care. The funder had no input in the writing of the manuscript or decision to submit for publication.

### Author Declarations

Ethical approval was not required for this analysis of secondary data

## References

1. Laverty AA, Diethelm P, Hopkinson NS, Watt HC, McKee M. Use and abuse of statistics in tobacco industry-funded research on standardised packaging. Tob Control [Internet]. 2015;24(5):422–4. Available from: http://tobaccocontrol.bmj.com/lookup/doi/10.1136/tobaccocontrol-2014-052051

2. Chantler (2014) Standardised packaging of tobacco - Report of the independent review undertaken by Sir Cyril Chantler (April 2014) https://webarchive.nationalarchives.gov.Uk/20140911094224/http://www.kcl.ac.uk/health/packaging-review.aspx.

3. World Trade Organization (2020). Australia — Certain Measures Concerning Trademarks, Geographical Indications and Other Plain Packaging Requirements Applicable to Tobacco Products and Packaging https://www.wto.org/english/tratop_e/dispu_e/cases_e/ds435_e.

4. Special Eurobarometer 482. Public perception of illicit tobacco trade [https://ec.europa.eu/commfrontoffice/publicopinion/index.cfm/Survey/getSurveyDetail/instruments/SPECIAL/surveyKy/2191].

5. Special Eurobarometer 443. Public perception of illicit tobacco trade [https://ec.europa.eu/commfrontoffice/publicopinion/index.cfm/Survey/getSurveyDetail/instruments/SPECIAL/surveyKy/2191].

6. Filippidis FT, Chang KKC, Blackmore I, Laverty AA. Prices and Illicit Trade of Cigarettes in the European Union: A Cross-sectional Analysis. Nicotine Tob Res [Internet]. 2020 Jan 8; Available from: https://doi.org/10.1093/ntr/ntaa004

7. van Walbeek C, Blecher E, Gilmore A, Ross H. Price and Tax Measures and Illicit Trade in the Framework Convention on Tobacco Control: What We Know and What Research Is Required. Nicotine Tob Res. 2013 Apr;15(4):767–76.

8. Gilmore AB, Rowell A, Gallus S, Lugo A, Joossens L, Sims M. Towards a greater understanding of the illicit tobacco trade in Europe: a review of the PMI funded ‘Project Star’ report. Tob Control [Internet]. 2013 Dec 11; Available from: http://tobaccocontrol.bmj.com/content/early/2013/12/ll/tobaccocontrol-2013-051240.abstract

